# Functional Connectivity Alterations in Asymptomatic Familial Early-Onset Alzheimer’s Disease Caused by APP Duplication

**DOI:** 10.1101/2024.09.25.24314362

**Authors:** Rotem Paz, Limor Kalfon, Eyal Bergmann, Ayelet Eran, Judith Aharon-Peretz, Tzipora C Falik Zaccai, Itamar Kahn

## Abstract

**BACKGROUND AND OBJECTIVES:** Autosomal dominant mutations of Alzheimer’s Disease (AD) are highly penetrant allowing to characterize the asymptomatic phase of this devastating condition. We investigated brain-wide functional alterations in the asymptomatic phase of autosomal dominant early-onset AD (ADEOAD) and explored whether a functional brain fingerprint of the disease can be found prior to clinical manifestation.

**METHODS:** In this cross-sectional study fourteen asymptomatic *APP*-dup carriers and eight *APP*-dup non-carriers from the same kindred underwent neurological and neuropsychological examination and resting-state fMRI scanning. The functional connectome of each participant was constructed based on 264 pre-defined brain regions, which were classified into seven different sensory (Visual and Somatomotor) and association (Default, Frontoparietal, Ventral Attention, Dorsal Attention, Limbic) cortical networks. Time courses were extracted from all regions in each participant and brain-wide Fisher’s *z*-transformed Pearson correlation (*z*(*r*)) from each region to all other regions was calculated, resulting in a 264 × 264 connectivity matrix per participant. These matrices were used for network similarity and connectome-based predictive modelling (CPM) analyses aimed to characterize age-dependent functional connectivity alterations within the carriers group that reflect their position along the neurodegeneration trajectory.

**RESULTS:** Comparing individual connectomes within the carriers group to the average connectome in the non-carrier group, we found that network similarity between groups decreased in an age-dependent manner. With age the APP-dup connectome diverged from the non-carriers connectome, with this difference being driven primarily by alterations in association networks and specifically involving the Default and Frontoparietal networks. Moreover, decreases in network similarity of association networks correlated with decreased performance on a visuospatial memory task. Using CPM, we found that in this family, carriers’ age can be predicted independent of the non-carriers control group and that this prediction is based mainly on connections within and between association networks.

**DISCUSSION:** Functional connectivity was used to assess the progress of AD pathology in asymptomatic carriers of an autosomal-dominant deterministic gene. Our results show that in asymptomatic *APP*-dup carriers, aging is associated with functional connectivity alterations that preferentially involve association networks. Understanding these alterations might lay the foundation for development of novel diagnostic markers and assist in determining the appropriate timing of therapeutic interventions in ADEOAD.

## 1. Introduction

Autosomal dominant early-onset Alzheimer’s disease (ADEOAD) is presently identified as linked either to point mutations or indels within one of the three amyloid processing genes: amyloid precursor protein (*APP*) on chromosome 21, and presenilin 1 (*PSEN1*) or presenilin 2 (*PSEN2*) on chromosomes 14 and 1 respectively^1^. Despite the fact that ADEOAD represents less than 1% of all AD cases^2,3^ it is well studied due to its high penetrance which allows to determine with high certainty the development of the disease. Moreover, ADEOAD and late onset AD share clinical and neuropathological characteristics including neuronal loss, neurofibrillary tangles, senile plaques, and cerebral amyloid angiopathy^4,5^. Therefore, ADEOAD may be especially well-suited to thoroughly characterize the asymptomatic phases of AD and generate models for prediction.

The use of neuroimaging, specifically functional magnetic resonance imaging (fMRI), in AD research may help to elucidate the nature of functional connectivity alterations during various phases of the disease. Resting-state fMRI (rs-fMRI) of AD patients consistently demonstrated decreased functional connectivity in the Default network (DN)^6–8^. In the asymptomatic phase of ADEOAD functional connectivity within both the posterior and anterior components of the Default network was consistently demonstrated to be disrupted^9–12^. Recently, more attention is drawn to studying connectivity in other high order cognitive networks (e.g., Frontoparietal and Ventral Attention networks)^6,13^ as well as using functional connectivity global measures of network integration and segregation to predict brain’s age^14^.

A commonly used method for estimation of functional connectivity is seed-based correlation analysis. This method relies on hypothesis-driven pairwise correlations and is thus limited as it is suboptimal for capturing global network attributes and higher-order interactions that drive functional connectivity^15,16^. To address this challenge data-driven methods that consider network-level effects were developed^17,18^. Another challenge in analyzing fMRI data is considering variability of functional connectivity measures across individuals. In the past, most neuroimaging studies collapsed data from many participants to detect differences in brain-behavior relations between groups of individuals. By doing so, these studies disregarded inter-subject variability within each group although, it has been recognized that brain functional organization varies between individuals^19–22^, and that this variability in functional connectivity is robust and reliable^19^. Moreover, it has been demonstrated that functional networks are dominated by stable individual factors, with much less than previously appreciated influence of task-state and day-to-day variability^20^, thus allowing to accurately identify individuals from a large group, serving as a functional "fingerprint"^19,20^. With the advent of these approaches, studying connectome individual variability and its association with individual brain organization and behavioral differences has become more prevalent^23,24^. Taken together, the aforementioned work highlights the importance of data analysis at the individual rather than the population-level and opens the door for their use in precision- and personalized-medicine approaches.

Here, we sought to examine whether individual whole-brain connectivity can indicate progression to disease onset among asymptomatic *APP*-dup carriers, and whether *APP*-dup carriers show a distinct pattern of whole-brain connectivity alterations compared to non-carriers family controls.

## 2. Materials and methods

### 2.1. Participants

Fourteen *APP*-dup carriers and eight *APP*-dup non-carriers from the same kindred were recruited for this study. Clinical characteristics of this cohort were previously described^25^. All participants were asymptomatic (did not report experiencing cognitive, behavioral, or functional deterioration before and during clinical data collection and did not meet diagnostic

criteria for MCI during neuropsychological/neurological evaluations). Participants were excluded if they had (a) lifelong history of delayed mental development, (b) current or past neurological disorders/injuries (e.g., TBI; stroke), and (c) current or past psychiatric disorders. Demographic data is presented **in Table 1**. The IRB committees of Rambam Health Care Campus, Galilee Medical Center and the Israeli Ministry of Health approved this study. Study participants signed informed consent forms. Participants received financial compensation for their participation (200 NIS) and a copy of their structural MRI scan.

**Table 1.** Demographic Data of rs-fMRI Participants.

|  | Non-Carriers (N=8) | APP-dup Carriers (N=14) |
| --- | --- | --- |
| Sex | 5 Females, 3 Males | 8 Females, 6 Males |
|  | Mean (Range) | Mean (Range) |
| Age | 36.38 (19-56) | 30 (19-39) |
| Years of Education | 12.88 (12-19) | 12.36 (11-14) |

### 2.2. Clinical and neuropsychological assessments

All participants were examined clinically by a neurologist and neuropsychologically by a neuropsychologist. Participants were assessed on (a) Global cognition tests: The Mini-Mental State Examination (MMSE); Montreal Cognitive Assessment (MoCA); (b) General intelligence (g) tests via four subtests from the Wechsler Adult Intelligence Scale-III (WAIS-III)^26^ battery: Similarities; Digit Span; Matrix Reasoning and Digit Symbol^27^; (c) Episodic memory tests: Paired Association Learning (PAL) from the CANTAB battery^28,29^, ‘What Was Where?’ task^30^, Verbal Paired Associates (VPA) subtest from the Wechsler Memory Scale-III (WMS-III)^31^; and (c) Psychological screening measures (Beck Depression Inventory – II (BDI-II)^32^, Beck Anxiety Inventory (BAI)^33^.

### 2.3. MRI data acquisition and preprocessing

Scans were acquired using a 3T scanner (type MR 750, version SIGNA 20, General Electric Medical systems America, Milwaukee, USA) and a 16 channels Head Neck Spine (HNS) Coil equipped with foam paddings to minimize head motion. T1 structural images were acquired using a spoiled gradient recall (SPGR) sequence (voxel size of 1 × 1 × 1 mm^3^, flip angle of 12°, matrix size of 256 × 256, number of slices was 156 and the field of view [FOV] was 25.6 × 25.6 cm^2^). Functional resting-state scans were done using a T2*-weighted gradient echo planar imaging (GE-EPI) sequence that allowed to evaluate the BOLD response. Participants were asked to remain with open eyes for two eight-minute scans (voxel size of 2.5 x 2.5 x 3.5 mm^3^, TR of 2500 ms, TE of 35 ms, flip angle of 90°, a matrix size of 128 × 128, FOV of 28.8 × 28.8 cm^2^) with 40 slices, and 200 volumes resulting.

### 2.4 Image preprocessing and quality assurance criteria

Preprocessing was performed following conventional methods^34,35^, using FSL (FMRIB Software Library v. 5.0.1, Oxford, UK) and SPM (Wellcome Department of Imaging Neuroscience, London, UK). The pipeline included removal of the first four volumes to allow for magnetic field stabilization, compensation for slice-dependent time shifts, rigid body correction for motion using a 6-parameter linear transformation, normalization to Montreal Neurological Institute (MNI) template (MNI152). Then, specific preprocessing for rs-fMRI was applied including removal of the voxel-wise mean signal and linear trend regression, regression of signal from ventricles, white matter and global average signal as well as motion estimates and their first derivatives, temporal band pass filtering (0.009–0.08 Hz), and spatial smoothing using a 6 mm full-width half-maximum Gaussian blur. Time-courses from regions of interest (ROIs) were produced from unsmoothed data. To correct for the impact of global motion and respiratory-related artifacts, we censored volumes with framewise displacement (FD) over 0.3 mm or the root mean square of the temporal change of the voxel-wise signal at each time point (DVARS) at 150% and interquartile range (IQR) above 75^th^ percentile.

### 2.5 Construction of the functional connectome

To construct the functional connectome of each participant, we used a well-established parcellation^36^ that includes 264 brain regions, which were classified into seven different cortical networks^37^, which was then divided qualitatively to sensory (Visual and Somatomotor) and association (Default, Frontoparietal, Ventral Attention, Dorsal Attention, Limbic) networks. After defining connectome nodes, we extracted their time courses in each participant and calculated the Fisher’s *z*-transformed Pearson correlation *r* values, resulting in a 264 × 264 connectivity matrix per participant. These matrices were used for network similarity and connectome-based predictive modelling analyses.

### 2.6 Similarity analysis

To characterize alterations in functional organization in asymptomatic *APP*-dup carriers, individual connectomes of the carriers’ group were compared to the average connectome of the non-carriers controls (**Figure 1A**). This comparison was conducted using the network similarity measure^20^, which was computed as Fisher’s *z*-transformed correlation between each *APP*-dup carriers connectivity matrix to the controls average connectivity matrix. Then, network similarity values derived from the whole-brain connectome or network specific versions of it, were compared to behavioral/demographic measures using Spearman’s correlation.

**Figure 1.**
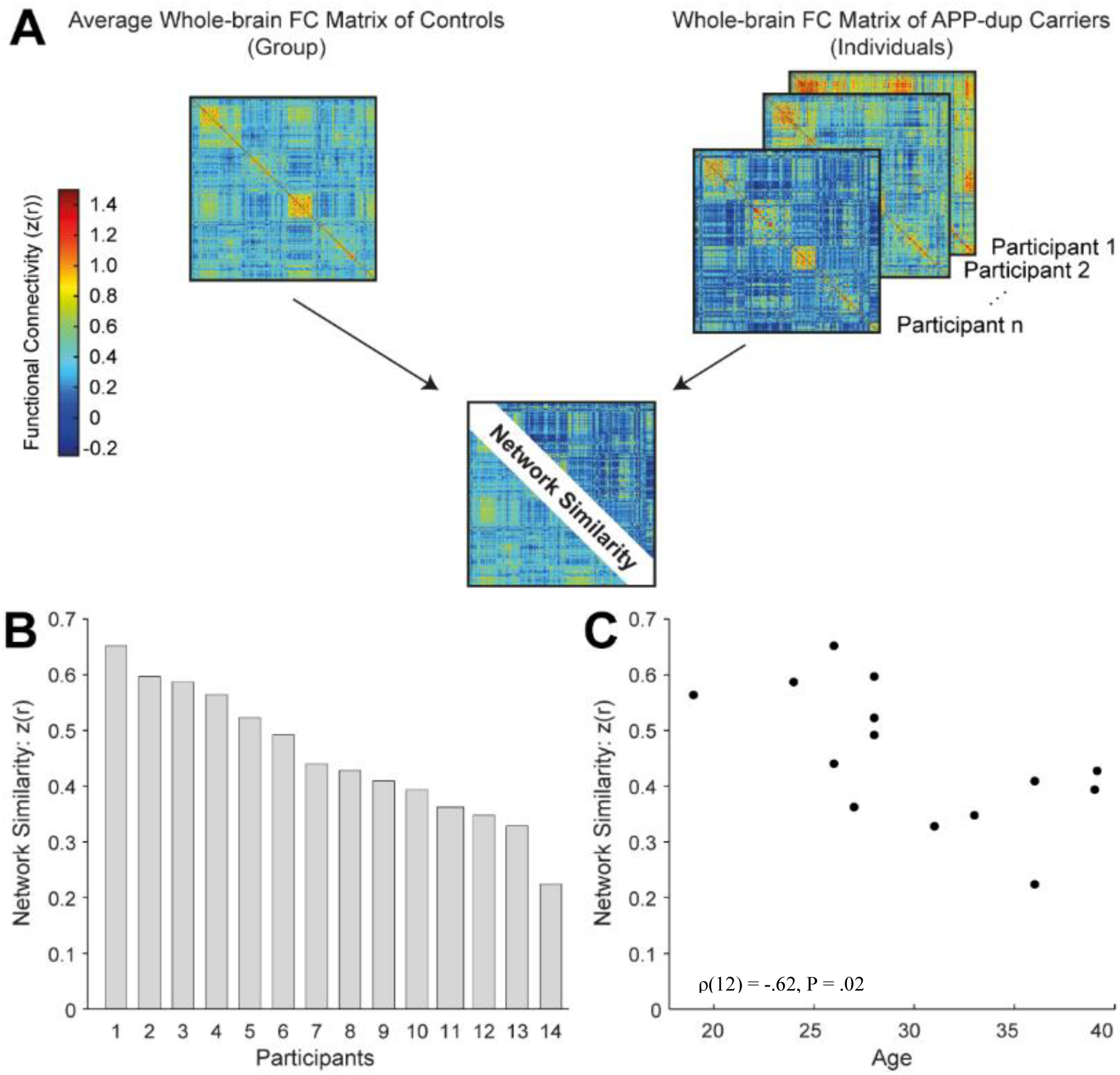
Individual Variation in *APP*-dup Carriers’ Functional Connectivity as Function of Age. **(A)** 14 *APP*-carriers’ whole-brain connectivity matrices were correlated to an averaged whole-brain connectivity matrix of 8 controls. Fisher’s *z*-transformed correlation value serves as a similarity index between *APP*-dup carriers and controls. **(B)** Individual variation of whole-brain connectivity similarity index in *APP*-dup carriers. **(C)** similarity index decreases as function of age. As participants age the *APP*-dup connectome diverge from controls’ connectome.

### 2.7 Connectome-based predictive modelling (CPM)

To link individual variation in the functional connectome to age variability, we followed a previously published detailed protocol for CPM^38^. For each participant, inputs to CPM were edges comprising whole-brain connectivity matrix and age. We used a leave-one-out cross-validation approach in which Spearman’s correlation was calculated between each edge in the connectivity matrices and age across *n* - 1 participants (i.e., *n* = 13) over 14 iterations. In each iteration, *p* < 0.01 (uncorrected for multiple comparisons) positive and negative correlations were selected and summarized to two values per participant, which were combined in a single linear regression model. This model was used for prediction of age in the *n*^th^ tested participant in each one of the 14 iterations. Finally, the correlation between predicted and observed age performance, which is statistically independent of the edge selection threshold, was calculated and formally tested by comparing it to the distribution of correlation in 1000 iterations in which age values were randomly assigned to participants. Since the model’s predictions are characterized by narrower distribution relative to age^38^, the predicted values were normalized. To characterize the edges that contributed to the prediction, we extracted the edges that were included in the model in all 14 iterations and presented them in matrix plots. Then, we examined whether the contributing edges were biased to connections between association or sensory regions using a set of Z-tests for independent proportions.

## 3. Results Similarity analysis

### Age-dependent reduction in network similarity in asymptomatic APP carriers

*APP*-dup carriers functional connectome diverges from non-carriers family controls. Using network similarity analysis, we found individual variation in the connectome of *APP*-dup carriers in comparison to controls (**Figure 1B**). Taking into consideration age being a proxy of disease onset (in the absence of Aβ/Tau burden measures and since we know the average age of clinical symptoms onset of this kindred), we hypothesized that this variation is related to the variable ages (age spectrum) in this group. Indeed, the whole-brain connectivity similarity index showed a significant negative correlation to age (ρ_(12)_ = -.62, *p* = .02). The older the carriers were, the lower their network similarity index to controls was (**Figure 1C**). To examine whether this effect stems from alterations in sensory and/or association networks, we calculated network similarity for two sub-matrices: Sensory (Somatomotor and Visual) and Association (Default, Frontoparietal, Ventral Attention, Dorsal Attention, Limbic). The similarity index for the Association networks showed a significant negative correlation to age (ρ_(12)_ = -.65, *p* = .01; **Figure 2A**), while similarity index for the Sensory networks and age were not significantly correlated (ρ_(12)_ = -.44, *p* = .12; **Figure 2B**). Taken together, these results suggest that association networks connectivity of carriers are changing and becoming less similar to those of controls as they age, while sensory networks connectivity are less affected.

**Figure 2.**
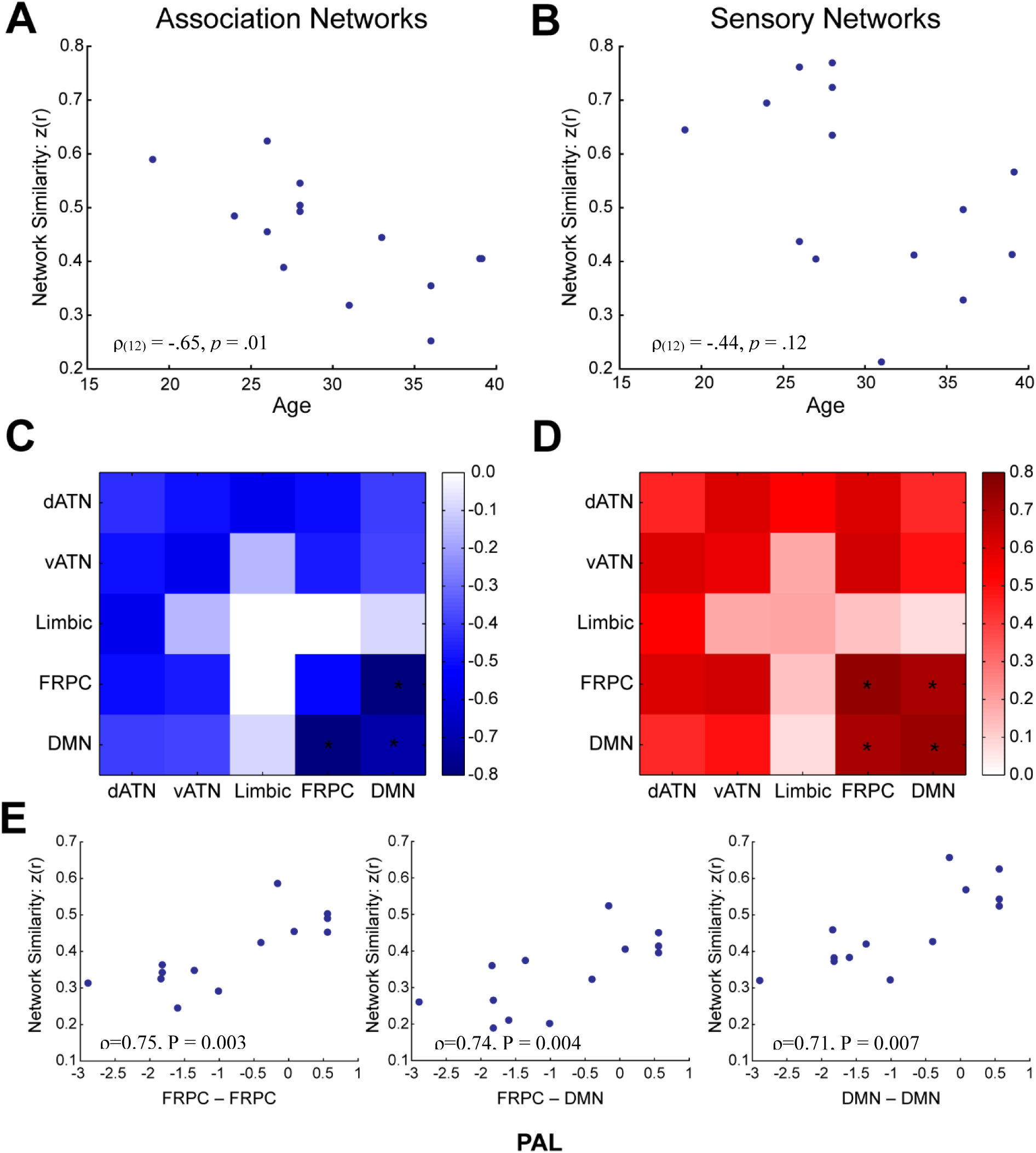
Alterations in Association Networks are Driving the Divergence of *APP*-dup Carriers’ Connectome from Controls’ Connectome, and their Performance on the PAL Task. (**A**) Similarity index for association networks significantly decreases as a function of age (ρ_(12)_ = -.65, *p* = .01). **(B)** Similarity index for sensory networks does not significantly change as a function of age (ρ_(12)_ = -.44, *p* =.12). **(C)** Correlations between functional connectivity within and between brain networks and age, showing that Inter-connectivity between the FRPC network and the DN, as well as the intra-connectivity within the DN are the main alterations correlated with age (* *p* < 0.05). **(D)** Correlations between functional connectivity within and between brain networks and performance in the PAL task showing that Intra-connectivity within the DN and within the FRPC network, as well as the inter-connectivity between the FRPC network and the DN is correlated with performance on the PAL task (* *p* < 0.05). **(E)** As the connectivity similarity index of these networks decreases, performance on the PAL task decreases and becomes impaired. dATN, dorsal attention; vATN, ventral attention; FRPC, Frontoparietal; DMN, Default.

To examine which networks are driving this effect, we restricted the analysis to specific connections within and between association networks. The inter-network connectivity between the Frontoparietal and the Default networks, as well as intra-network connectivity within the Default were significant (*p* = 0.009 and *p* = 0.03, respectively; **Figure 2C**). To examine whether the alterations we found in the connectivity of association networks is correlated with the participants’ performance on the PAL test, a test that assesses visuo-spatial memory and new learning. This analysis included 13 *APP*-dup carriers and 8 controls who had this behavioral result available. The similarity index for each network to itself and to all other association networks was then correlated to the PAL 6 shapes total errors measure (6 shapes stage being the highest stage all participant managed to be tested on). Results showed that the similarity indices of the Frontoparietal and Default inter-network connectivity were correlated with PAL (ρ = 0.71, *p* = 0.007), and so was the similarity index of the intra-network connectivity of the Frontoparietal (ρ = 0.75, *p* = 0.003) and the Default (ρ=0.74, *p* = 0.004; **Figure 2D**). This result was specific to the 6 shapes stage, suggesting that this brain-behavior association emerges only under cognitive burden. The more similar the connectivity between carriers and non-carriers was, the better the performance on the PAL was (**Figure 2E**).

### Connectome-based predictive modeling (CPM)

A similarity index may change due to a consistent change in a specific network across all subjects or due to alterations in different networks between participants. To address this idiosyncratic nature of similarity index, we sought to build a connectome-based predictive model (CPM)^38^ to link between functional connectivity profiles in a leave-one-out cross-validation (LOOCV) analysis, in which the age of an *APP*-dup carrier is predicted based on connectivity patterns and their correlation to age in the rest of this group. CPM demonstrated good correspondence between predicted and observed age (*r*(12) = 0.775; **Figure 3A**). To formally test the goodness of prediction, we shuffled the age data 1000 times and ran LOOCV analyses on shuffled data to extract significance level (*p* = 0.001), which confirmed significant prediction. Importantly, age values were not correlated with individual average head motion estimates (mean frame displacement) during scanning (r(12) = 0.16, *p* = 0.58), confirming that this result is not artifactually introduced by motion. Finally, we found that while both positive and negative correlations between and within association networks contribute to the prediction of age in this model, most contributing edges were positive (219 out of 254, 86.22%). Examining the distribution of these edges between networks, we found that positive edges contributed to the model predominantly (74.73%) were inter-network edges (**Figure 3B**). Moreover, we found that the Frontoparietal, Ventral Attention and Default inter-network connectivity contained the main positive correlations contributing to this model, and that the Default-Frontoparietal and Dorsal Attention-Frontoparietal inter-network connectivity, as well as inter-network connectivity within the Default contained the main negative correlations contributing to this model. To formally test this observation, we used a set of two-tailed *Z*-tests for independent proportions (**Figure 3C**) to test whether significant edges are biased toward connections between Association nodes (A:A, n = 70/14331), Sensory nodes (S:S, n = 26/12464) or one Sensory and one Association node (S:A, n = 10/4970, all comparisons were corrected for multiple comparisons using the Bonferroni method). This analysis confirms that both positive and negative correlations are more frequent in A:A connections than in S:A (Z = 3.83, *p* < 0.001, corrected) and S:S (Z = 2.72, *p* = 0.02, corrected) connections with no bias toward S:A relative to A:A connections (Z=-0.097, *p* = .92, corrected).

**Figure 3.**
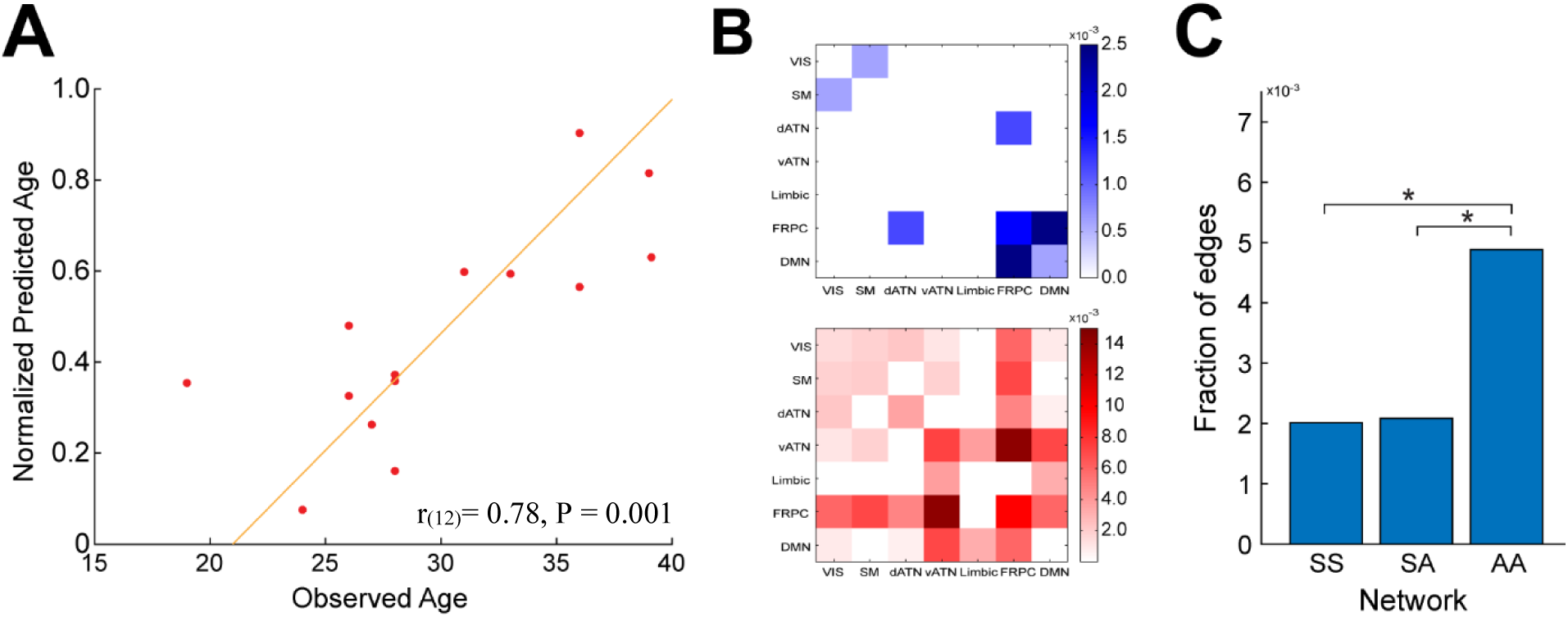
Connectome-based predictive modeling of individual’s Age. **(A)** Observed and predicted age (n=14 *APP*-dup carriers) are highly correlated. A formal statistical analysis was done using a shuffling analysis in which connectome-based predictive modeling was applied to shuffled age data, the original r-value designates the first highest value out of 1000 iterations. **(B)** Representations of edges that were negatively (top-blue) and positively (bottom-red) correlated with age in all leave-one-out cross-validation iterations. **(C)** All edges from the previous analysis were divided to three categories: connections between two association regions (A:A, n=70), connections between an association and a sensory region (S:A, n=10) and connections between two sensory regions (S:S, n=26). The results revealed that association-association edges are most predictive of age.

## Discussion

Our results show that the connectome of *APP*-dup carriers diverges from this of non-carriers controls in an age-dependent manner already at the asymptomatic stage. This divergence is seen approximately around the age of 30, preceding the conversion from asymptomatic to symptomatic phases by approximately 10-20 years. Age-dependent alterations preferentially involve association networks, and specifically the Default and Frontoparietal networks. Altered connectivity patterns between and within these networks are correlated to visuospatial memory performance, and specifically to the ability to generate associations. The more different the connectivity between *APP*-dup carriers and non-carriers controls was, the worse the performance on the PAL visuospatial memory task was. To control for a potential idiosyncratic divergence that might be captured by the similarity analysis and to avoid the dependency on the control group, we then computed a CPM model. The model predicted for each *APP*-dup carrier their brain-based age which is a proxy for their position along the neurodegeneration trajectory. Analysis of the edges contributing to prediction revealed that connections from association networks and specifically the Default, Frontoparietal and Ventral Attention, contributed the most to the model. Collectively, these results indicate that alterations in functional connectivity precede cognitive decline in APP-dup carriers, and that if these changes are directly related to neuropathological processes, then they will localize predominantly to association regions in the Default and Frontoparietal networks.

The main motivation for the current study was to identify neural correlates of neurodegeneration at asymptomatic phases of ADEOAD. This type of research, better known as brain-wide association study (BWAS), is widely used in cognitive and clinical neuroscience. A seminal study by Marek et al.^39^ has recently shown that in the general population BWAS requires thousands of participants, with lower sample sizes resulting in unreproducible associations. Following this seminal observation, it was suggested that clinical populations with larger effect sizes might be used for BWAS in much smaller cohorts^40^. Here we followed this approach by examining the neural correlates of ADEOAD, a condition with dramatic effects on cognitive performance and the brain, which strongly depend on age. In addition, several other elements in the study design increase the sensitivity of the analysis. First, compared to studies in the general population, an examination of a single kindred provides better control for environmental and genetic factors that affect brain architecture^41^. This advantage is important for BWAS, as demographic variability was shown the be associated with predictive model failures.^42^ Moreover, we used multivariate functional measures, which are superior to structural or univariate measures^39^. A possible explanation is that a multivariate approach captures network-level alterations, which are less variable than region-level changes in the context of brain pathology^43^. Nevertheless, future studies of this mutation and other ADEOAD are needed to validate the current result and examine the extent of generalizability of the reported alterations.

In the context of AD, functional connectivity studies have uncovered alterations with disease progression^44–46^. Recent application of computational models demonstrated successful prediction of brain age based on neuroimaging data. Millar et al.^47^ demonstrated that functional connectivity can identify variations across adulthood and effectively forecast brain age among amyloid-negative, preclinical late-onset sporadic AD, and symptomatic late-onset sporadic AD participants. In their model apparent brain age was quantified and compared to normative neuroimaging trajectories. Their findings may suggest a biphasic response to preclinical AD pathology where individuals in the preclinical phase show “younger” patterns of FC, and individuals in the symptomatic phase show “older” patterns of FC. This is perhaps a reflection of compensatory response to early AD pathology, allowing these individuals to maintain normal cognition despite accumulating pathology as previously suggested^48^. In contrast, Gonneaud et al.^14^ have shown that in ADEOAD the biological progression of AD is characterized by an advanced pattern of brain aging detectable before symptom onset, particularly in individuals with rare genetic mutations causing AD and significant Aβ pathology. Our results are in line with those of Gonneaud et al.^14^. Indeed, we observe that asymptomatic individuals with strong genetic predisposition exhibit a distinct pattern of functional brain changes in a monophasic fashion. The CPM model allowed us to determine the existence of a linear trajectory of deterioration as a function of age.

In our study, age also demonstrated a behavioral relevance, so that an “older” FC pattern within the APP-dup carriers correlated with decreased performance on the PAL 6 shapes memory task. Since age serves as a proxy of neurodegeneration in our cohort, it will be interesting to examine its relationship to biological measures of neurodegeneration. Specifically, since our cohort is a group of individuals with amyloid beta (Aβ) overproduction, we hypothesize that as age progresses, Aβ accumulates. Future studies including Aβ/Tau-PET examination will allow to further elucidate the connections between Aβ/Tau aggregation, resting-state fMRI functional connectivity, structural alterations, and behavior in this unique group.

Examining which networks showed age-dependent alterations in asymptomatic carriers revealed the Default and Frontoparietal networks were predominantly affected. Importantly, these networks are related to emotional processing, self-referential mental activity, recollection of prior information and experiences, cognitive control and decision-making processes^49,50^, all of which are domains that are affected in AD. Second, Badhwar et al.^7^ suggested in their metanalysis a strong tendency in the literature toward specific examination of the Default network. Our results demonstrate that functional connectivity alterations go well beyond this specific network.

Last, a major limitation of the current study is the use of cross-sectional analyses. Longitudinal examination of this unique cohort would be useful to address intraindividual reliability and the trajectory of brain age estimates as AD pathology progresses over time. Such analysis can better address questions regarding resilience and susceptibility among APP-dup carriers. In **Figure 3A** there are several examples of individuals who are at the same chronological age but differ as to their predicted brain age based on their functional connectivity. Currently, we cannot determine if this variability is clinically relevant, and specifically if this difference in the connectome predicts earlier onset of AD. A longitudinal follow up will allow to explore implementation of this model for precision medicine purposes, and to develop a method for predicting deterioration based on the specific individual functional connectivity aging trajectory.

## AUTHOR CONTRIBUTIONS

Rotem Paz: Initiated the project with IK, JAP, designed the models and tasks, conducted neuropsychological assessments, acquired the fMRI data, preprocessed the data, conducted statistical analysis, interpreted the results, and co-wrote the paper.

Limor Kalfon: coordinated all aspects of genetic evaluation of the project and co-wrote the paper.

Eyal Bergmann: designed the models, conducted statistical analysis, interpreted the results, and co-wrote the paper.

Ayelet Eran: evaluated clinically all MRI scans.

Judith Aharon-Peretz: Initiated the project, participated in patient recruitment, performed physical and neurological examinations, reviewed the paper.

Tzipora C. Falik Zaccai: Initiated the project, supervised and participated in recruitment of participants. Supervised and designed the genetic aspect of the project. Analyzed the genetic findings and reviewed the paper.

Itamar Kahn: Initiated the project, co-wrote, reviewed and finalized the paper. All authors revised and approved the manuscript.

## Data Availability

All data produced in the present study are available upon reasonable request to the authors

## Acknowledgements

First, we would like to thank the participants of this study. We would like to thank the MRI unit staff at Rambam Health Care Campus, Haifa, Israel: Dr. Anat Greenfeld, Ms. Diana Dana, Ms. Hana Vakrat, Ms. Sabina Sendler, Mr. Yosef Tal-Dan, Mr. Amir Gaber, Ms. Ola Einav, Ms. Ella Goltzman, Ms. Rozet Hija. Finally, we would like to thank Dr. Daniela Lichtman for helping with figures preparation.

## Funding

This study was supported by a research grant from TEVA Pharmaceutical Industries Ltd. and the Lowell R. Lamb Research Fund for Alzheimer’s Diseases (Grant No. ATS 11393).

## Notes

### Competing Interest Statement

The authors have declared no competing interest.

### Author Declarations

The IRB committees of Rambam Health Care Campus, Galilee Medical Center and the Israeli Ministry of Health approved this study.

### Summary of Updates

The first name of the author Judith Aharon-Peretz is misspelled in the submission information and two stylistic issues were corrected in the abstract.

